# Curve-fitting approach for COVID-19 data and its physical background

**DOI:** 10.1101/2020.07.02.20144899

**Authors:** Yoshiro Nishimoto, Kenichi Inoue

## Abstract

Forecast of the peak-out and settling timing of COVID-19 at an early stage should help the people how to cope with the situation. Curve-fitting method with an asymmetric log-normal function has been applied to daily confirmed cases data in various countries. Most of the curve-fitting could show good forecasts, while the reason has not been clearly shown. The *K* value has recently been proposed which can provide good reasoning of curve-fitting mechanism by corresponding a long and steep slope on the *K* curve with fitting stability. Since *K* can be expressed by a time differential of logarithmic total cases, the physical background of the above correspondence was discussed in terms of the growth rate in epidemic entropy.

## 1. Introduction

COVID-19 outbreak was first reported in Wuhan, China, then the disease soon spread to Europe, US and worldwide in the first half of 2020. Japanese government declared a state of emergency in April and people were requested to stay at home. The number of daily cases was the biggest concern and people were anxious to know the possible timing of the peak-out and settling because they have been threatened not just by the deadly disease but also by its critical impact on the economic activities. Our trial approach is to keep people informed of the macroscopic trend of cases which should help them to think of their future.

Among epidemic analysis methods the SIR theory has provided a reliable mathematical model^1)^, however, it requires not only infected (I) data but also suspicious (S) and recovered/died (R) data to evaluate an actual ‘reproduction number’^2)^. Those data may include time lag and it is not regarded as a real time method. Although PCR test data include a weekly periodicity and strong arbitrariness, macroscopic trends could be extracted if the arbitrariness remain consistent. Therefore curve-fitting approaches could be a simple forecast tool even though it is kind of phenomenological one depending also upon people’s activities. T. Tomie proposed a curve-fitting with a normal distribution to answer the real time request and applied it to COVID-19 in Wuhan^3)^ since it was known that flu epidemic followed a normal distribution^4)^. It offered an informative forecast, however, the model required ‘ad hoc’ ideas of different parameters for increase/decrease phases because the COVID-19 profile has a noticeable feature to leave a trail in an asymmetric manner.

We applied a curve-fitting with a log-normal distribution to meet the asymmetric features. This paper describes the curve-fitting approach and its physical background as specifically as possible in order for people to follow it as a simulation tool for their own countries/regions and to relieve people’s concern on COVID-19.

## 2. Curve-fitting

The fitting was carried out in the following manner.

1. Database: daily cases by country provided by CSSE at Johns Hopkins University ^5)^
2. Model: log-normal function for daily cases A log-normal function expressed by formula (1) has an asymmetrical distribution in Fig.1.

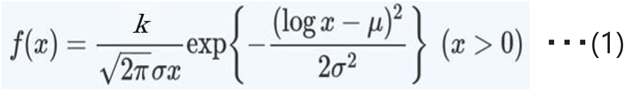 A log-normal appears in the distribution of incubation periods of viruses^6)^.
3. Fitting: using solver in Excel add-on (GRG nonlinear search for least mean square) An example of spread sheet templates for a curve-fitting computation is shown in Fig.2. If you input daily cases data, forecasting date and initial values for undetermined coefficients a, b, c of the log-normal function in the yellow cells, Excel’s solver searches for an optimal coefficients’ combination to minimize the variance Σ(n(t)-f(t))^2^ less than the threshold level. The curve-fitting result is depicted in the right-hand graphs. The upper shows the fitting with daily cases and the lower shows the cumulative distribution to fit the total cases. Thus curve-fitting can be carried out in the straight forward manner without any ad hoc techniques.

**Fig.1.**
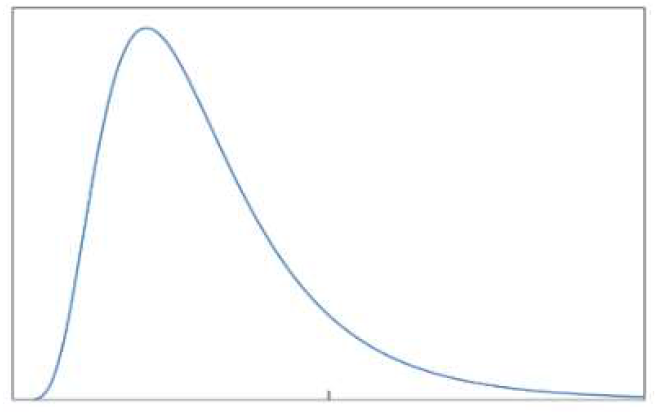
Log-normal function

**Fig.2.**
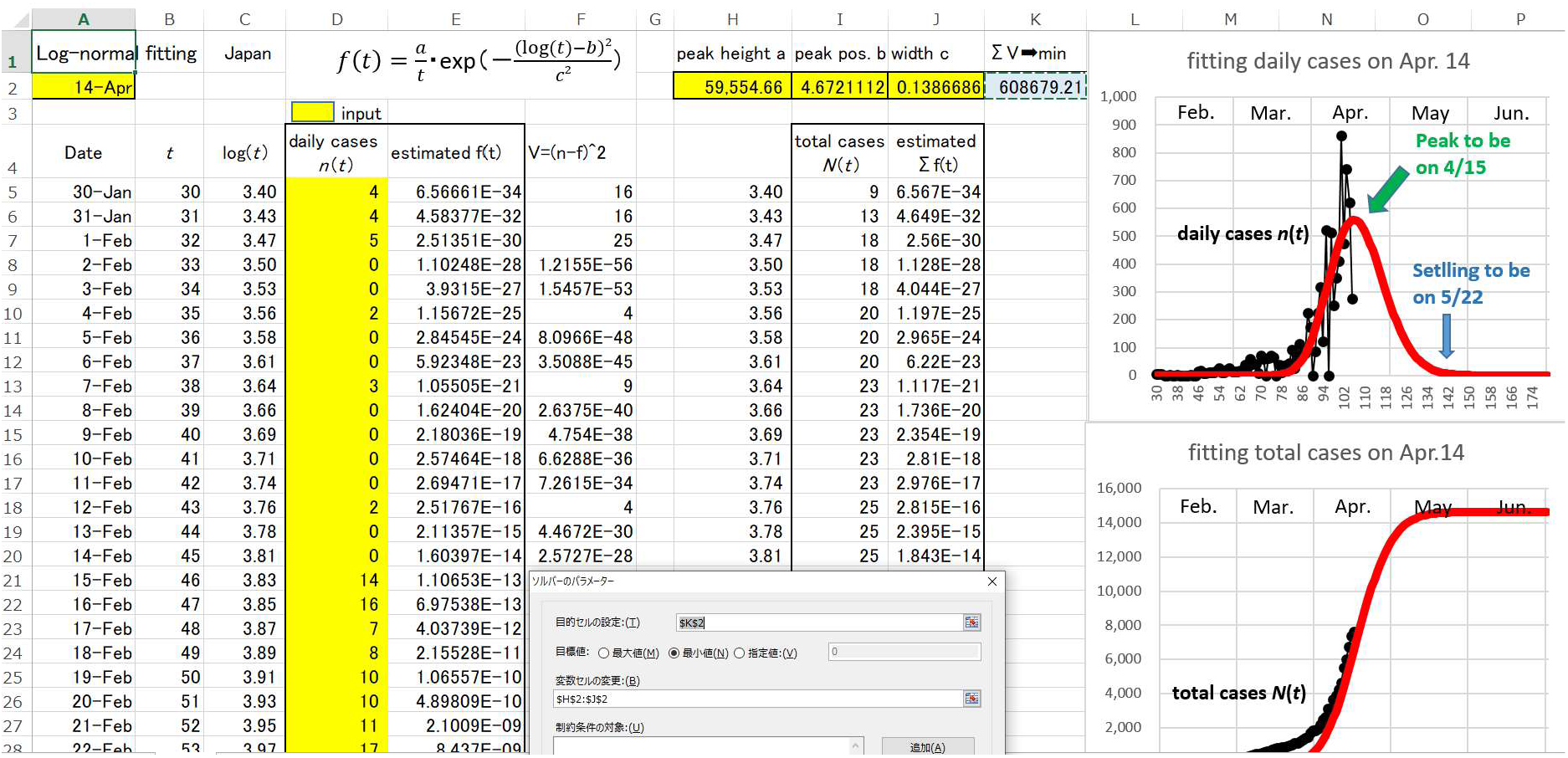
An example of spread sheet template for curve-fitting

Log-normal function fittings with daily cases in Japan were carried out since early April. Although Excel solver could not find optimal coefficients until Apr. 11^th^, it suddenly became successful on the next day. Since then we repeated fitting periodically, among which typical examples are shown in Fig. 3 for April 12^th^, 16^th^, 20^th^, 25^th^, May 5^th^ and 25^th^, respectively. The horizontal axis is the date number originated by Jan. ^1st^. ‘Settling’ in the graphs shows a date expected that daily cases go below the 1/100 of peak cases along the fitted curve (in red). The forecast on Apr. 12^th^ looks a little bit pessimistic, however it is getting reliable as days go by.

**Fig.3.**
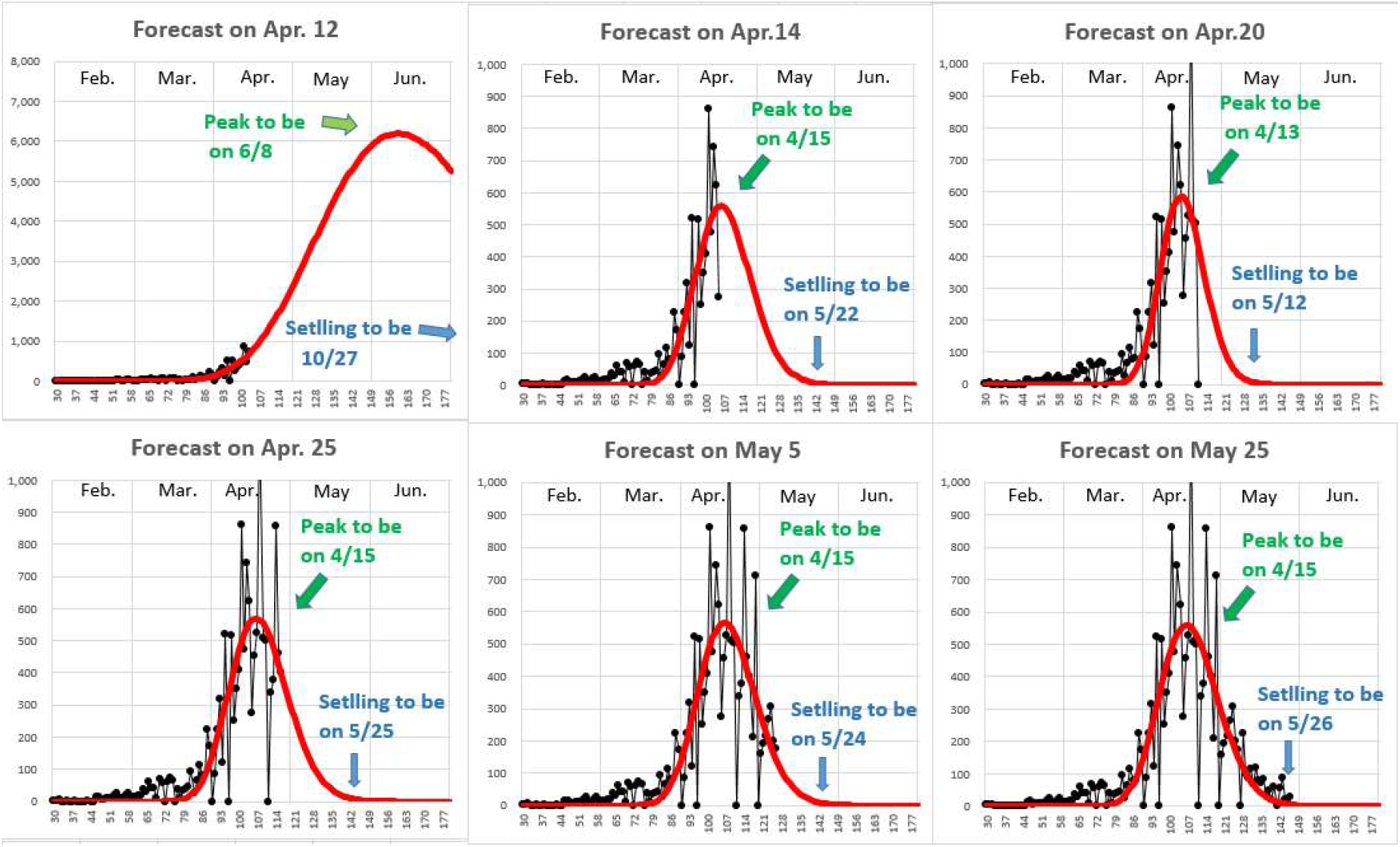
Forecast using curve-fitting with daily cases in Japan at various dates

These series of graphs in Fig.3 show that significant curve-fittings are possible a few days after Apr.12^th^ in the case of Japan and forecast of peak-out and settling date is getting firmer day by day as depicted in Fig.4.

**Fig.4.**
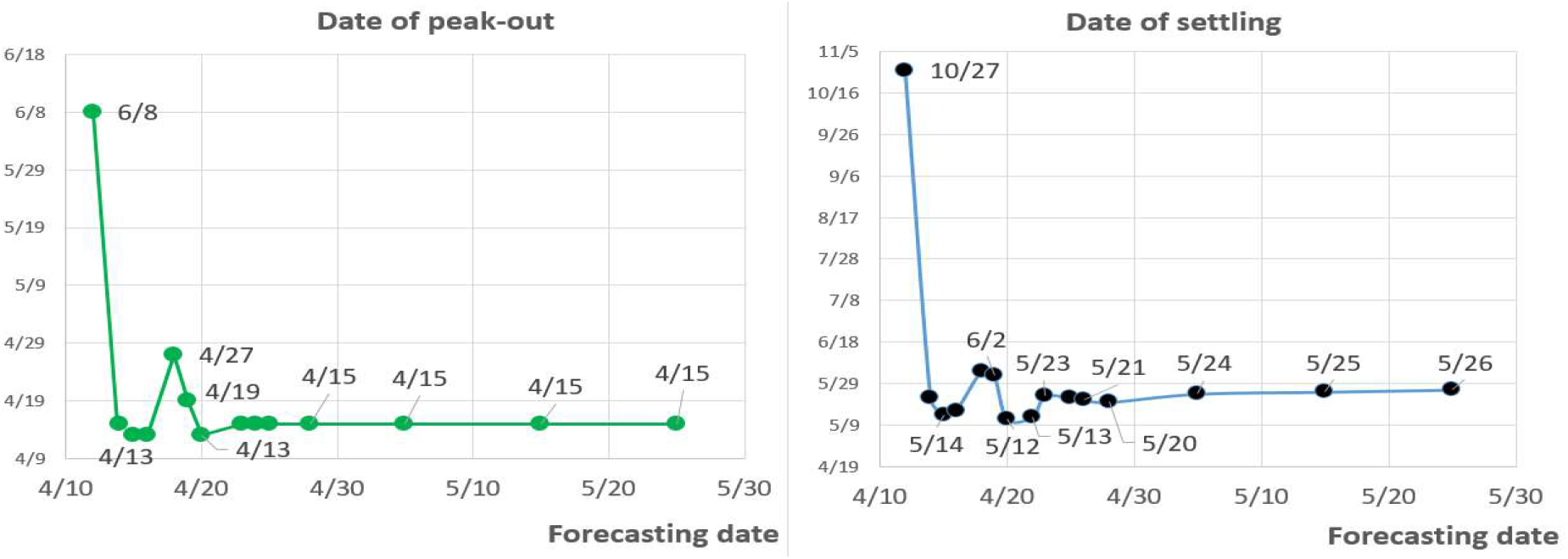
Forecast of peak-out and settling date

The forecast peak-out date is so consistent that there is no question that daily cases reached a peak on Apr. 15^th^ in Japan. On the other hand, a state of emergency was declared on Apr.7^th^ at seven selected areas and on Apr. 16^th^ across Japan, and the government strongly requested the public to stay at home to avoid 80% human contact. The effect of the government request would have come out two weeks later around Apr. 22^nd^ at earliest.

Consequently the daily confirmed cases in Japan had already peaked out one week prior to the timing when the effect of such emergency action were expected. Was the declaration of emergency state given too late or even not necessary? It was understandable that Japanese had too much self-restraint because of the very first experience. However, they have to verify the cost/merit in case of upcoming waves of COVID-19 because self-restraint in the whole society will cause huge economic damages and losses.

By the way why was the curve-fitting successful after Apr. 12^th^? We had no reasoning for it and could not answer the question, just saying ‘please ask the data’.

## 3. Reasoning of curve-fitting

Why and when does the curve-fitting get successful in the course of epidemic? Are there any reasons for it? We had been thinking about the question and finally came across the *K* value^7)^, which was proposed by T. Nakano and Y. Ikeda and is defined by

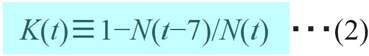

where *t* is the number of days from the reference date, and *N*(*t*) and *N*(*t*−7) are the total cases on days *t* and (*t*−7), respectively.

The relationship between curve-fitting on the daily cases and the *K* value in Japan is described in Fig. 6. The *K* value shows a long downward slope since the *K’*s summit dated Apr. 12^th^ while the curve-fitting was getting stable since the same date.

**Fig.5.**
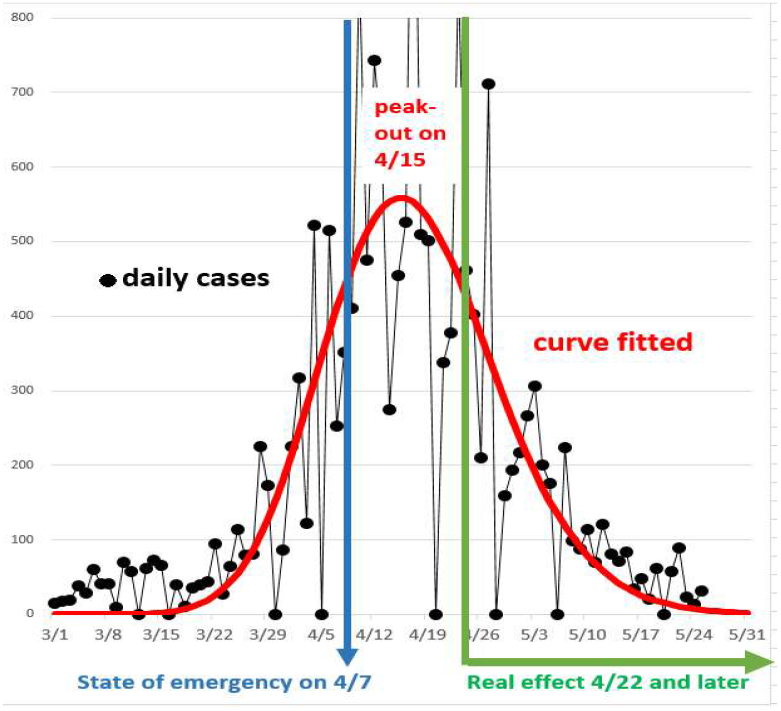
Relation between epidemic peak and effect of emergency state

**Fig.6.**
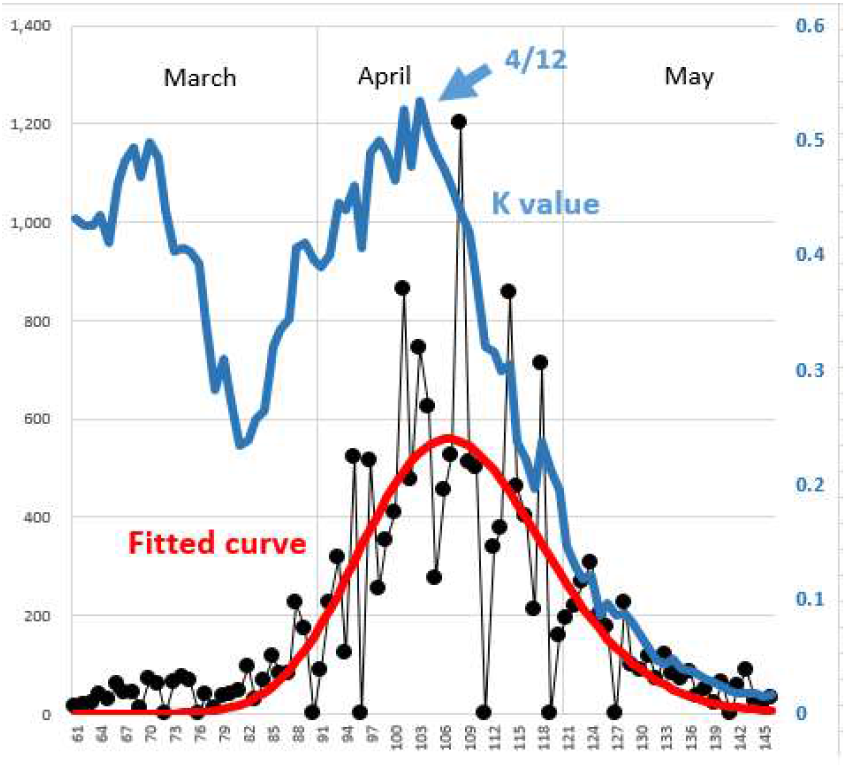
*K* value and fitted curve in Japan

Next we looked into the same relationship in other countries. Since USA total included multiple waves one after another, we chose New York as a representative. The *K* value graphs show long and steep slopes for Italy, Germany, New York and Taiwan in Fig.7 and curve-fitting was getting stable along the slopes initiated from the *K’*s summits similarly with the case of Japan. However, most profiles of the daily cases left trails as they were getting close to settling stages and the forecast of settling date got shifted later.

**Fig.7.**
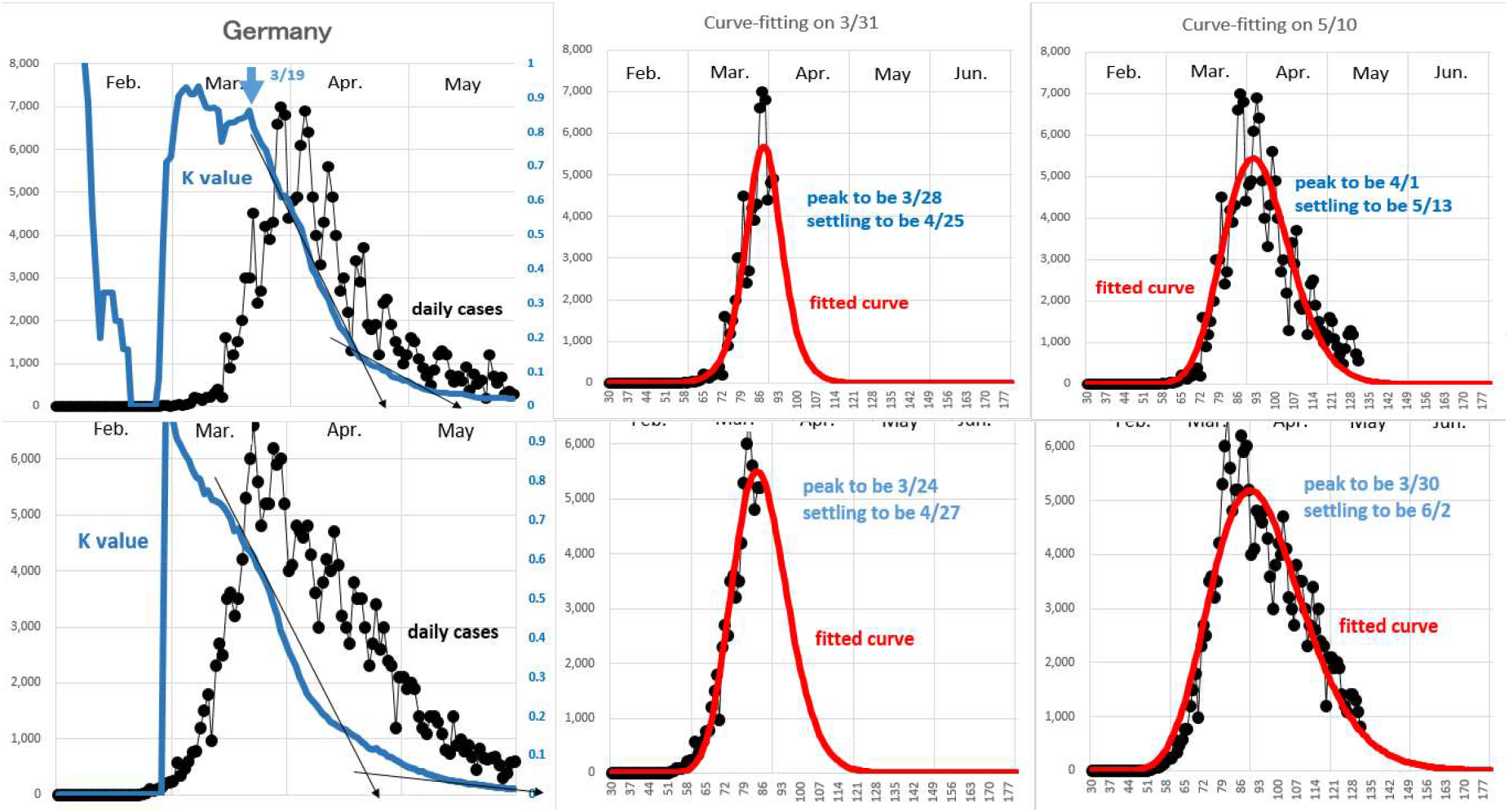
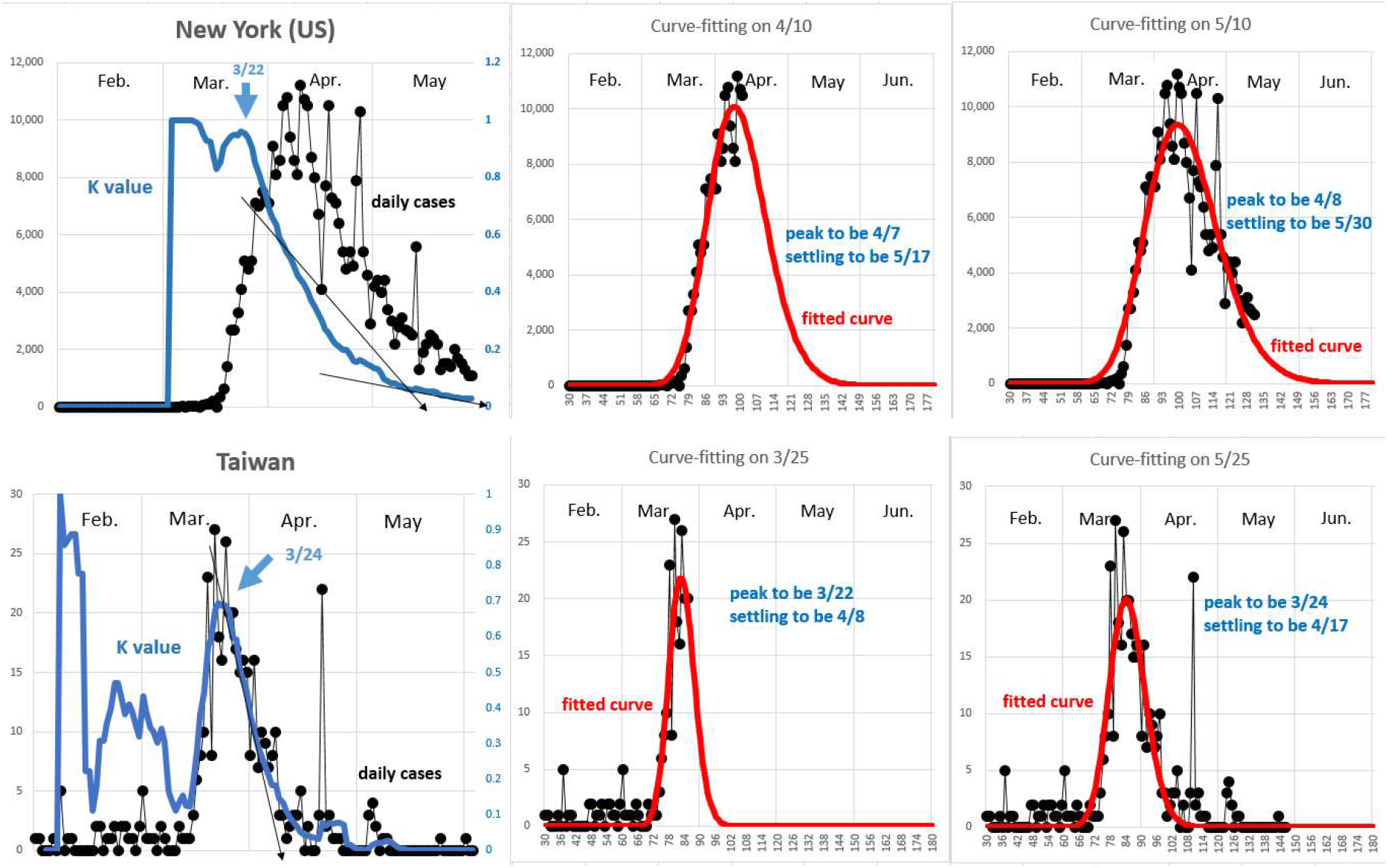
*K* values and fitted curves for Italy, Germany, New York (US) and Taiwan

In Sweden and Russia daily cases profiles show lingering patterns depicted in Fig.8. At first the *K* curves show steep slopes down to landing floors but gentle slopes are followed from the landing floors. Log-normal functions still fitted the Russian and Swedish data but the settling period might be quite volatile and delayed to late summer.

**Fig.8.**
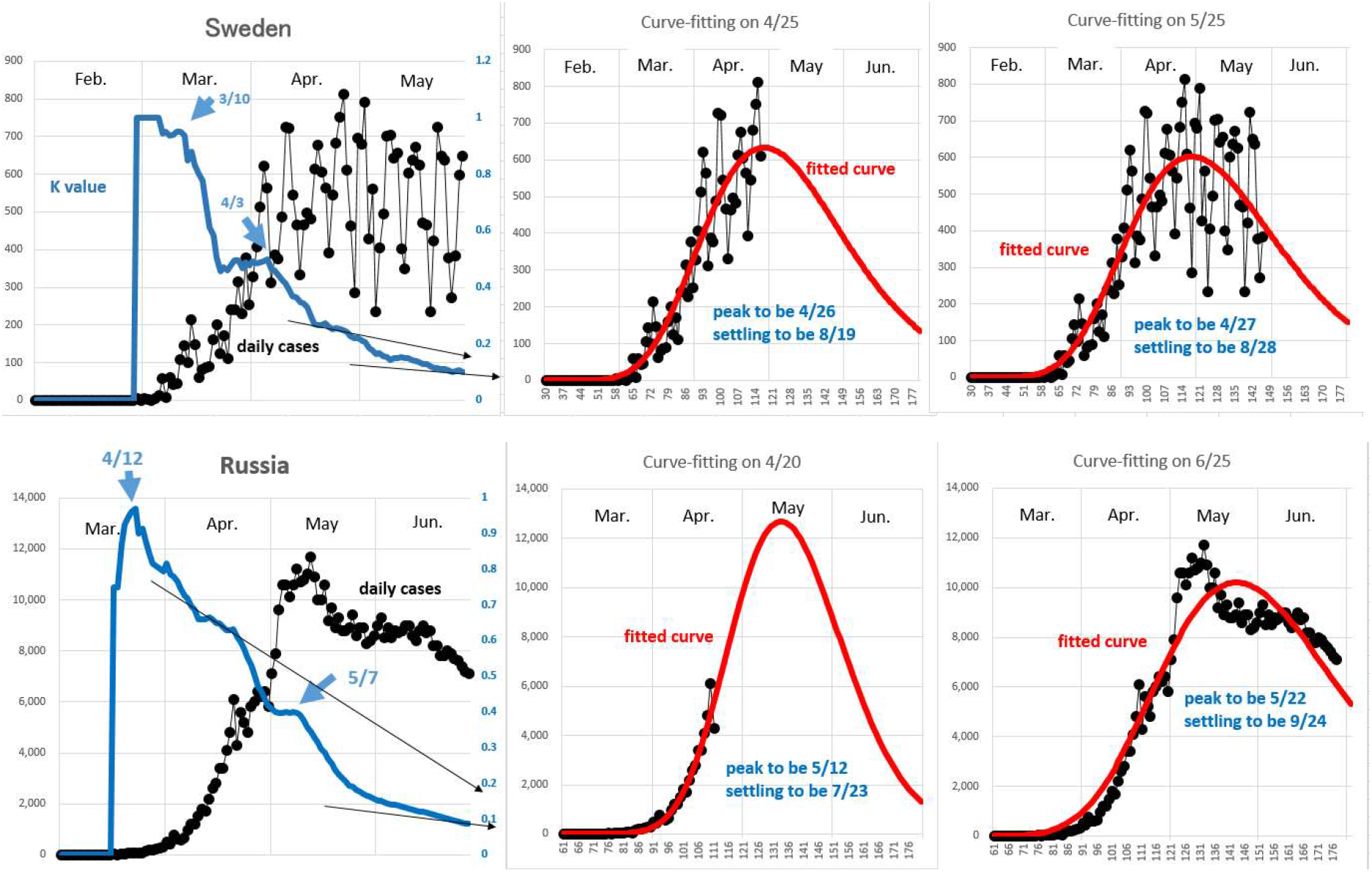
*K* value and fitted curve for Sweden and Russia

On the other hand, India, Brazil and South American countries are still suffering from exponential growth and even a sign of peak-out has not appeared yet. The *K* curves show gentle downward slopes with small ups and downs, which leads to an unstable curve-fitting and consequently to uncertain forecast of peak-out or settling periods in Fig.9

**Fig.9.**
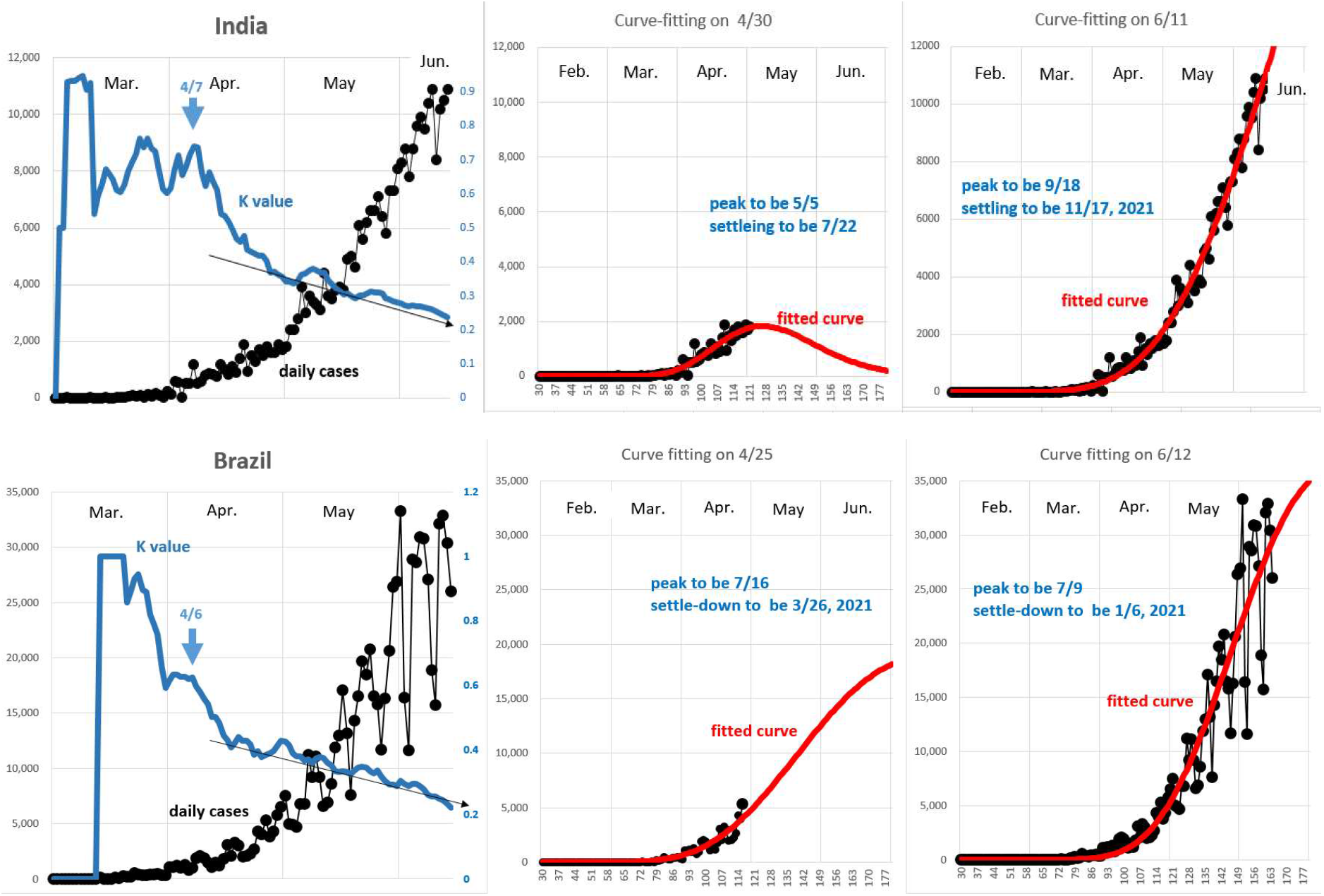
*K* values and fitted curves for India and Brazil

Let us summarize the daily cases profile, *K* value at the initial, peak-out, settling and *K’*s slope and curve-fitting stability in Table 1.

**Table 1.** The correlation between the K value and curve-fitting stability

| | start-to-peak<br>(months) | peak-to-settle<br>(months) | $K$ value at | | | $K$'s slope | Curve-fitting<br>stability |
| --- | --- | --- | --- | --- | --- | --- | --- |
|  |  |  | initial | peak-out | settling |  |  |
| Japan | 1 | 1.5 | 0.54 | 0.5 | 0.02 | steep | stable |
| Germany | 1 | 1.5 | 0.86 | 0.5 | 0.03 | steep | stable |
| Italy | 1 | 2 | 1.00 | 0.4 | 0.01 | steep | stable |
| NY(US) | 1 | 2 | 0.96 | 0.4 | 0.01 | steep | stable |
| Taiwan | 0.5 | 1 | 0.67 | 0.6 | 0.03 | steep | stable |
| Sweden | 2 | 4+ | 0.95⇒0.50 | 0.2 | (TBD) | gentle | case by case |
| Russia | 2 | 4+ | 0.97⇒0.40 | 0.2 | (TBD) | gentle | case by case |
| India | 3 | 5+ | 0.74 | <0.2 | (TBD) | gentle | unstable so far |
| Brazil | 3 | 5+ | 0.89⇒0.63 | <0.2 | (TBD) | gentle | unstable so far |

It was confirmed that the curve-fitting method had a considerably good stability even before the peak-out stage once the *K* curve entered to a steep slope in spite of noisy data. However, if we apply it to a forecasting tool to inform people of the epidemic outline from an outbreak through a peak-out up to a settling stage, we have to investigate the reliability of the tool carefully. There are several factors which could affect the reliability of the curve-fitting forecast although the background of them is still in a fog;

1. the accuracy estimated by the *K* curve
2. the variation caused by an asymmetric lingering effect
3. the effect of *K* value at initial in Japan and Taiwan

## 4. Discussions

Now let us discuss on the factors which affect the forecasting reliability of the curve-fitting not only from the *K* curve geometry but from the physical meaning of the *K* value.

(1) The accuracy estimated by the *K* curve

In the previous chapter we confirmed that the fitting gets much firmer as the *K* curve has a steeper slope. This correlation is understandable from the *K* curve geometry. For example, the epidemic settling date can be estimated by an extension line along the *K* slope as shown in Fig.7∼9 and the date estimation accuracy Δ*t* is approximately given by

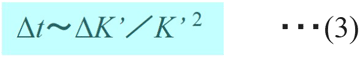

where *K’* is a time differential of *K* which means the gradient of the *K* slope. It is quite reasonable that a long (small Δ*K*’) and steep (large|*K*’|) slope on the *K* curve makes a small Δ*t*, that leads to a reliable forecast.

(2) The variation caused by an asymmetric lingering effect

As shown in Table 1, most of the countries’ COVID-19 data have peak-to-settling time much longer than start-to-peak one, that is, an asymmetric profile. This is the main reason why we have applied a log-normal distribution as the model function. Correspondingly the *K’*s slope has a trend of getting gentle and gentle as epidemic is getting close to settling. This spoils the reliability of forecasting of the settling period although the reliability of forecasting of the peak-out is not so much damaged by the lingering effect. What is the cause behind the asymmetric profile and the corresponding gentle slope in the *K* curve?

Asymmetries frequently appeared in the COVID-19 daily cases data have generally been explained with political interventions such as the lock-downs and quarantines in order to urgently suppress upcoming spread peaks and to avoid the medical care breakdown.

However, the asymmetries more or less occurs in most countries independent from the scale of the political intervention. For example, the lingering effect in Sweden data remarkably lasted more than two months although they have a unique policy of an early acquiring of ‘herd immunity’ with a less intervention.

(3) The effect of *K* value at initial in Japan and Taiwan

Generally speaking, if the *K* value at initial is sufficiently high, the slope in the *K* curve gets steeper and the peak-to-settling time gets shorter. Why does Japan have a short peak-to-settling in spite of its low *K* value at initial?

We have no idea about (2) and (3) so far and let us discuss on them from a physical background of the *K* value if possible. Let us remind of the definition of the *K* value;

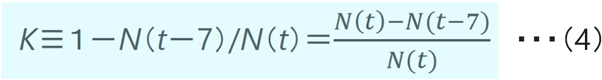

where *N*(*t*)-*N*(*t*-7) in the numerator is the total cases during seven days from day (*t*-6) to day *t*, that is,

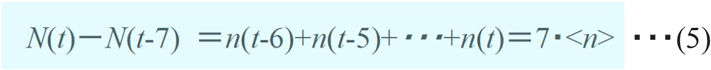

where *n*(*t*) is the daily cases of on day *t* and <*n*> is a moving average of *n*(*t*) during the previous week. Although *n*(*t*) generally changes volatilely, <*n*> shows a smoother profile. Here, note that the total cases *N*(*t*) and the daily cases *n*(*t*) have the following relationship;

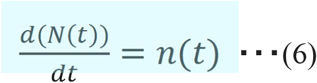

Using formula (5) and (6), The *K* value can be expressed in a different way as shown in the following formula as long as *n*(*t*) is not zero.

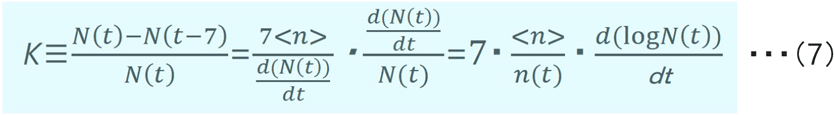

Thus the *K* value can be written with two factors <*n*>/*n*(*t*), a time-variant ratio, and *d*(log*N*(*t*))/*dt*, a time differential of logarithm of the total cases *N*(*t*). The profiles of the two factors are shown for Italy, Germany and India in Fig.10. The upper graphs show the actual data and the lower show the ones free of deviations based upon fitted curves.

**Fig.10.**
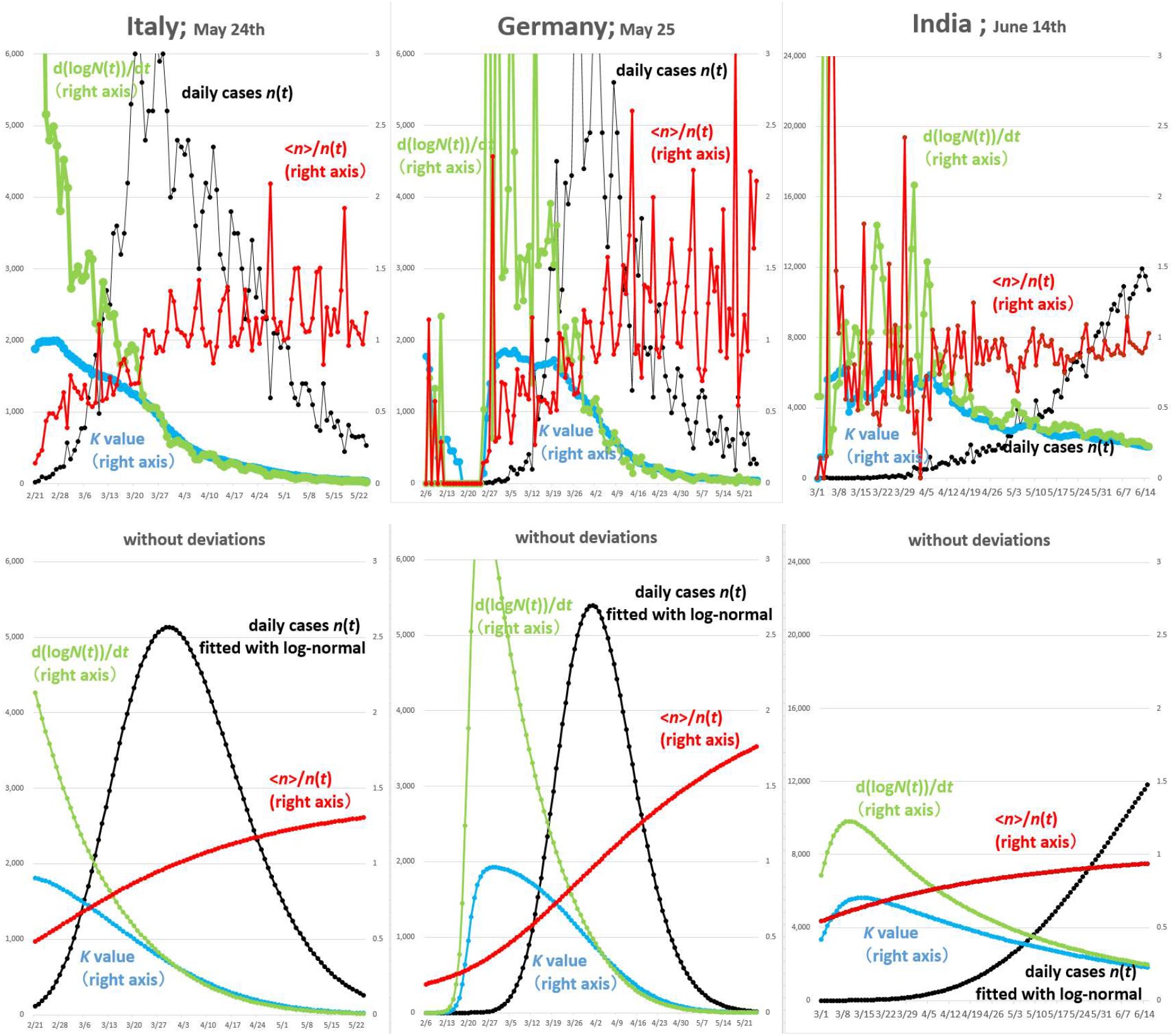
*K* value and its two factors for Italy, Germany and India

A time-variant <*n*>/*n*(*t*) is a small positive number informing a peak-out when it gets across value one. In this context the peak-out in India does not seem to be so far away. On the other hand what is the meaning of the factor *d*(log*N*(*t*))/*dt* ? We have been looking at graphs named ‘Logarithmic (of *N*(*t*))’ on the dashboard shown in Fig.11 in the COVID-19 database of CSSE at Johns Hopkins University to find a first sign of spread settling. It reminds us that *d*(log*N*(*t*))/*dt* has a certain relationship with an epidemic settling.

**Fig.11.**
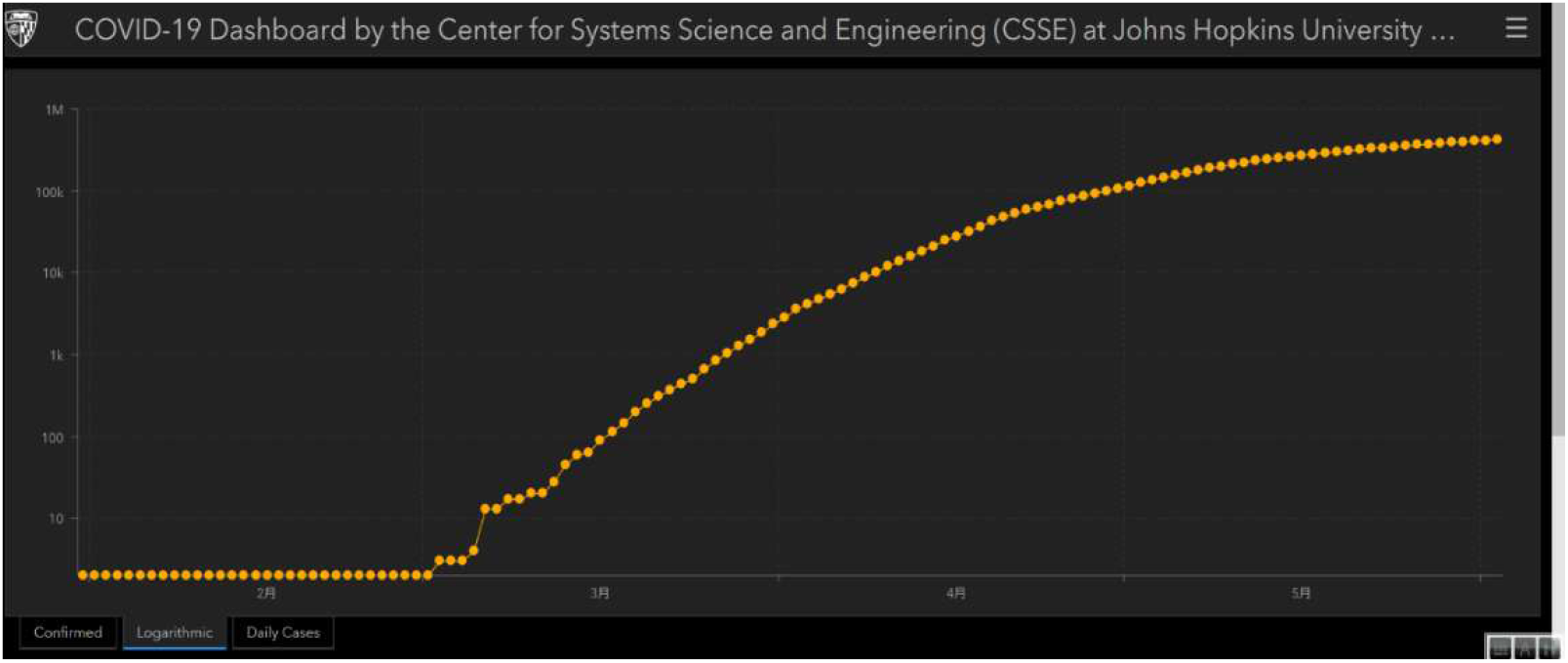
Example of logarithmic graph in database at Johns Hopkins Univ. (Source: Russian data on June 1^st^, 2020)

Let us consider on the meaning of log*N*(*t*) in the formula (7). As described in the supplementary materials, a total cases *N*(*t*) is well fitted with a logistic function. Incidentally the Fermi-Dirac distribution in the quantum statistics has exactly the same functional form with a logistic function although the variables, ‘time’ and ‘energy’ are different with each other. Fermi–Dirac statistics describe a distribution of many identical particles that obey the Pauli exclusion principle when the systems reach a thermal equilibrium, that is, the maximum entropy. It is reasoned by analogy that a total cases *N*(*t*) expressed with the similar logistic function has a certain relationship with a process of entropy growth under a certain physical constraints. If we assume that logarithm of total cases *N*(*t*) is an entropy *S*(*t*) in an epidemic process, the *K* value expressed in (7) can be written by the following formula;

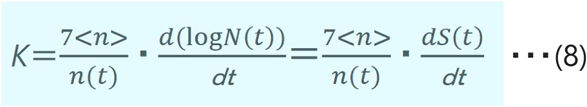

Since the time variant ratio <*n*>/*n*(*t*) is a positive number around one, the *K* value can be recognized as an entropy growth rate in the epidemic.

Therefore it is concluded that epidemic settling is an approach to a maximum entropy after spread. In the statistical physics a virtual ‘Maxwell’s Damon’ is trying to microscopically discriminate and sort different kinds of particles in order to keep the local entropy lower. Correspondingly, world countries are now trying to macroscopically reduce their economic activities through cities’ lock-down and isolation of positives as if they were keeping the local epidemic entropy lower.

Now that we understand the *K* value is a good estimate of the epidemic entropy growth rate, let us be back again to the questions (2) and (3) discussed above.

(2) What is the cause behind the asymmetric profile?

In other words why does a *K* curve change direction to escape zero when it is getting close to zero? From the entropy-based discussion above, *K*=0 means that the epidemic entropy *S* stops increasing and the local system has already reached a thermal equilibrium. However if any human interventions are given, the equilibrium will be surely disturbed and another entropy growth will get restarted. The lingering pattern of daily cases and an asymmetric distribution is worldly common and more or less an unavoidable matter because of human intervention. Forecasting with curve-fitting unavoidably depends upon human activities.

(3) Why is *K* value at initial strangely low in Japan and Taiwan?

Since the *K* value can be regarded as the epidemic entropy growth rate, this means that the epidemic entropy in Japan and also in Taiwan started growing quite slowly along the rate of doubled/7 days as schematically shown in Fig. 12 ^8)^. According to the statistical physics, the entropy growth rate generally depends upon physical conditions such as the density, pressure, strength of interaction and, of course, temperature of the local system. It can be inferred by analogy that Japanese weak interaction derived from their life style, always keeping clean with an appropriate physical/social distance, contributes to the initially low entropy growth rate even though the population densities in Japan and especially in Tokyo are considerably high. Or are there any reasons for the low entropy growth rate in Japan, owing to another ‘factor x’^2)^?

**Fig.12.**
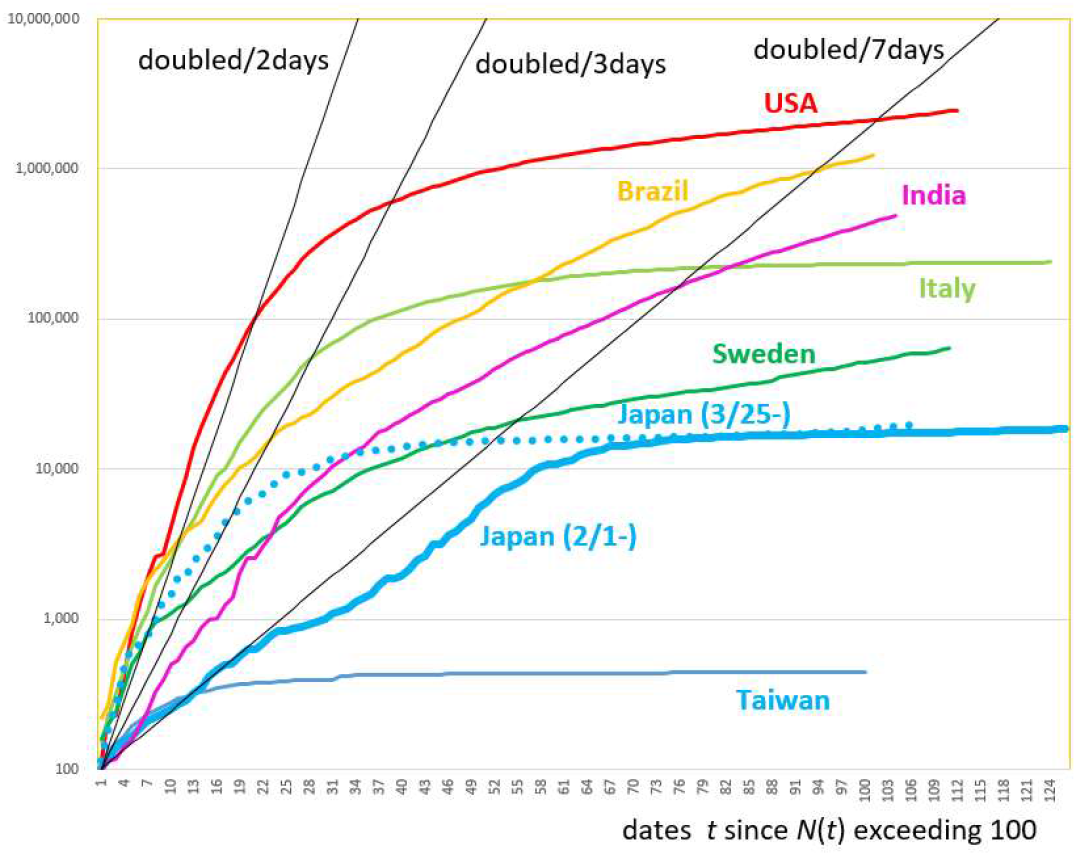
Comparison diagram of log *N*

T. Nakano explained that the low *K* value in Japan was caused by one of Wuhan strains which was prevalent in advance in the mid-March 2020 ^9)^. If the corresponding daily cases were taken away from Japanese data, the initial *K* value was not necessarily irregular as shown by the dotted line in Fig. 12. Further discussions are highly appreciated.

## 5. Conclusion

Daily given COVID-19 related news continued to depress many people and we believed that keeping the people informed of the macroscopic trend of confirmed cases should help them to set up their future plan.

We applied a curve-fitting approach using an asymmetric log-normal as a model function. Daily cases data in many countries were successfully fitted and timings of peak-out and possible settling could be forecasted at rather earlier stages. The reason why and when curve-fitting was possible was supported by the *K* value as a guideline.

The physical meaning behind the relationship between the curve-fitting approach and the *K* value was discussed in terms of epidemic entropy. The lingering effect inherent to COVID-19 and the difference in the initial *K* value was also discussed.

Under huge social and economic sacrifices, each government is now struggling on the containment of spread by using its brake and accelerator and will be studying on ‘how each country’s policy contributed or disturbed’ and ‘what was the cost/merit’. We hope that the review with the curve-fitting method and the viewpoint of epidemic entropy will play a role in the research review and, as well, in detecting the second wave which is very likely to flare up anywhere in the world.

## Data Availability

COVID-19 dashboard CSSE Johns Hopkins Univ

https://coronavirus.jhu.edu/map.html

## Acknowledgements

The authors thank Dr. T. Tomie for his original curve-fitting idea, Dr. Y.Yamaguchi for his timely suggestion on log-normal function, Prof. T. Nakano for his useful advices on the *K* value and Dr. Y. Kawata and Mr. Y.Yokota for constructive discussions.

## Supplementary materials

Let us consider on the meaning of log*N*(*t*) in the formula (7). In the previous chapter a log-normal function was adopted to fit data of daily cases *n*(*t*). However in order to fit data of total cases *N*(*t*), a logistic function seems suitable. A logistic function defined by formula (S1) shows step-like curves depicted in Fig.S1.

**Fig.S1.**
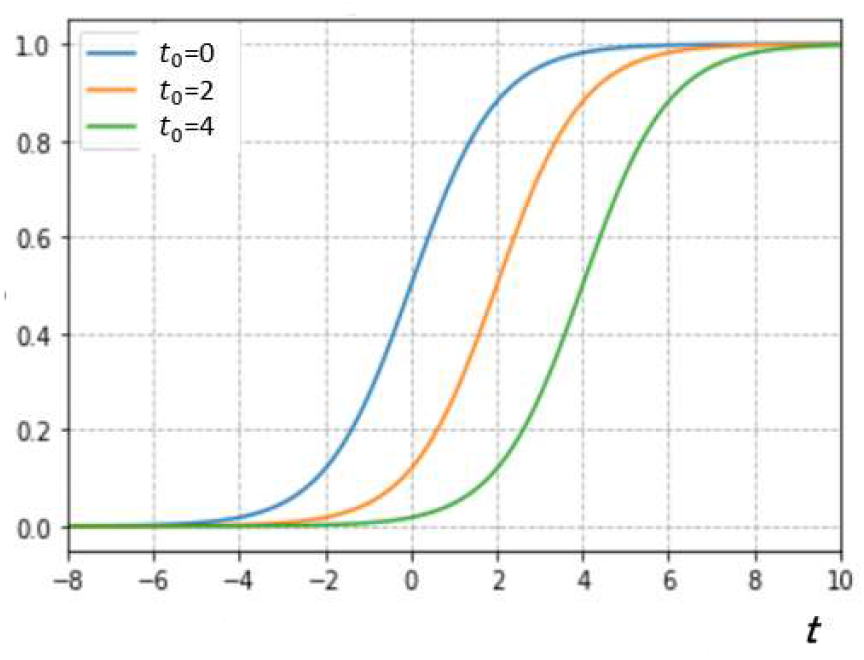
Logistic function

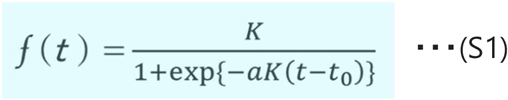

This function has been used for analyses in the world population increase, growing process of species and also epidemic spread under the environmental constraint. Actually the total cases of COVID-19 in Japan can be accurately fitted using a logistic function as shown in Fig.S2. Fig.S3 depicts the time differential of the logistic function in Fig.S2. It can be seen that the corresponding daily cases in Japan is followed as long as a symmetric distribution.

**Fig.S2.**
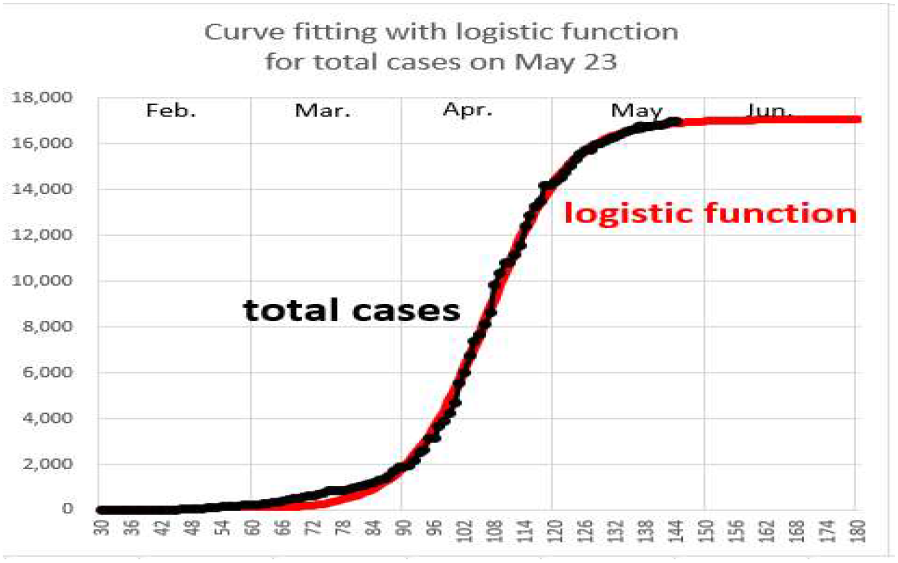
Total cases fitted with logistic funct.

**Fig.S3.**
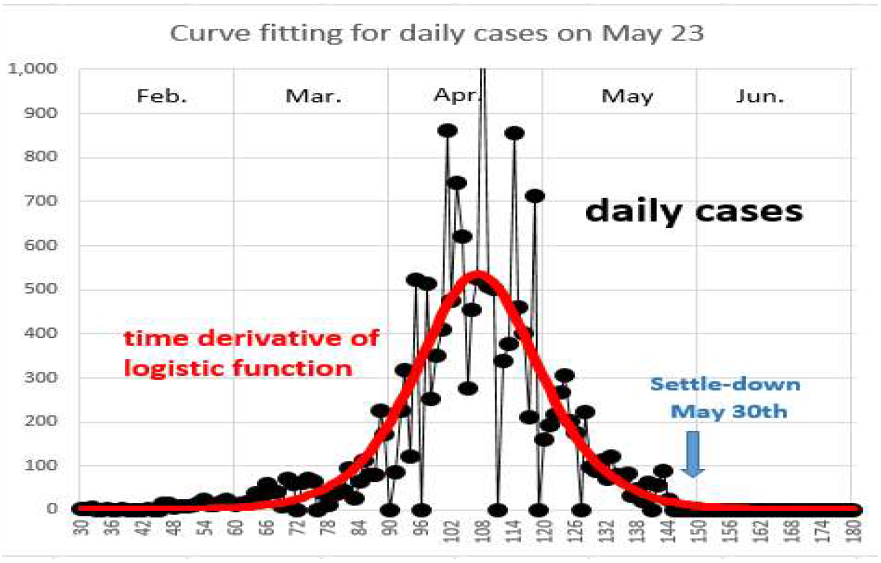
Daily cases fitted with logistic funct.

Therefore it can be concluded that the total cases *N*(*t*) is well-fitted with logistic functions. The expression (S1) reminds us of Fermi-Dirac distribution in the quantum statistics which. is given by a logistic function depicted in the formula (S2) and Fig.18, where β is an inverse temperature, ε is an energy of the single-particle, and μ is the total chemical potential.

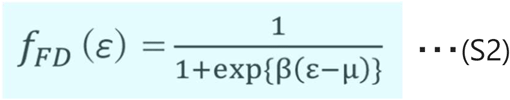

**Fig.S4.**
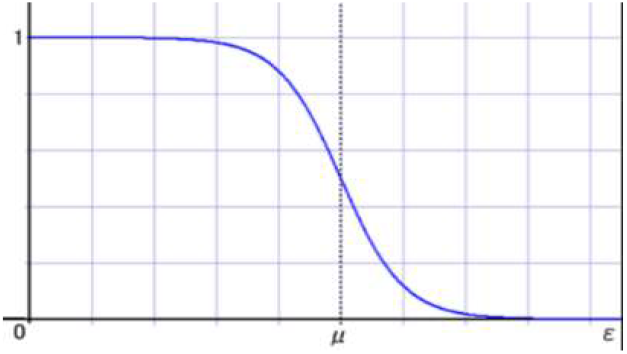
Fermi-Dirac distribution

Let us compare the formula (S2) with (S1). Although the variables, ‘time *t*’ in (S1) and ‘energy ε’ in (S2), are different with each other, functional forms are exactly the same and distributions are actually very similar. Fermi–Dirac statistics describe a distribution of many identical particles that obey the Pauli exclusion principle when the systems reach a thermal equilibrium, that is, the maximum entropy. It is naturally suggested that logistic function has a certain relationship with a process of increasing entropy under some physical constraints. If we assume that logarithm of total cases *N*(*t*) is an entropy *S*(*t*) in an epidemic process, the *K* value expressed in (7) can be written by the following formula;

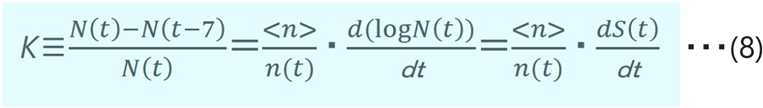

(EOD)

## Notes

### Competing Interest Statement

The authors have declared no competing interest.

### Funding Statement

none

